# Advancing the Safe Motherhood Initiative: a qualitative and sentiment analysis of local physician’s perspectives on antibiotic self-medication during pregnancy in a low- and middle-income country

**DOI:** 10.1101/2025.05.30.25328658

**Authors:** K Umeh, S Adaji, M Sacks, GU Eleje, EO Umeh, S Ushie, CG Okafor, CB Oguejiofor, U Bawa, S Bature, N H Madugu, R Singh, H Karuppuchamy

## Abstract

Although the Safe Motherhood Initiative is currently a global priority, the implications of maternal self-medication for meeting Safe Motherhood and Sustainable Development Goal 3 objectives in low- and middle-income countries has yet to be addressed. Although local medical doctors are an influential stakeholder group, able to determine health policy, how they view and feel about the problem of antimicrobial self-medication during pregnancy is not well understood. Thus, this study explores physicians’ views and sentiments regarding antibiotic self-medication in pregnant women from a West African country. We used mixed qualitative and quantitative analytic approaches. Semi-structured interviews were conducted with 25 medical doctors working in three tertiary hospitals. Thematic analysis was employed to identify key perspectives, while sentiment analysis was used to determine the emotional tone, based on an open-source pre-trained machine learning model for natural language processing. Several checks for methodological rigour were performed, including reviewing records of over 800 email conversations, and conducting respondent validation. Seven distinct themes emerged depicting views on antimicrobial treatment (e.g., easy availability of antibiotics), patient behaviour (e.g., use of medicinal herbs) and policy guidelines on antibiotics stewardship (lack of clear protocols). The prevailing sentiment portrayed a predominantly neutral demeanour towards antibiotic self-medication during pregnancy (χ2 (1, N = 1484) = 1314.858, *p* < 0.001), with an unusually high number of neutral labels, compared with positive (z = −36.058, *p* < 0.001) and negative (z = −36.410, *p* < 0.001) categories. The results of this investigation can provide useful information for managing antibiotic self-medication in pregnant women from resource-deprived regions where medical doctors are influential stakeholders. Our findings can be used to tailor local Safe motherhood policy initiatives on antimicrobial stewardship during pregnancy such that they address physician’s concerns and sentiments, including insufficient clinical practice guidelines and an ostensible lack of urgency.

## Introduction

In 1987 the World Bank, in collaboration with World Health Organisation (WHO) and United Nations Population Fund (UNFPA), launched the Safe Motherhood Initiative (SMI) to help raise global awareness about the impact of maternal mortality and morbidity (1). The primary goal of SMI is to achieve a significant reduction in maternal mortality (2). To achieve this goal the United Nation’s (UN) 193 Member States launched Sustainable Development Goals (SDGs) at a major summit in September 2015. SDG-3, which aims to ensure healthy lives and promote well-being for all, includes specific targets for maternal, neonatal and child mortality (3, 4). Although the SMI has been a global priority in recent years, the implications of maternal self-medication (i.e., using non-prescribed drugs in pregnancy) on meeting SDG-3 initiatives in low- and middle-income countries (LMICs) have yet to be addressed.

Self-medication with antibiotics is a world-wide problem (5–9). It is a particular concern during pregnancy (10, 11), especially in LMICs, where use of medication without a doctor’s prescription is a common practice. Over 50% of antibiotics consumed in developing countries involve self-medication (e.g., purchased over the counter) (12). A systematic review of 13 published articles on self-medication in pregnancy (N = 6202 pregnant women) found an overall prevalence rate of 32%, with one in five women (20.9%) self-medicating with antibiotics (10). Maternal self-medication is particularly problematic LMIC, with the highest prevalence rates found in Nigeria (72.4%) (10). In addition to readily available over-the-counter drugs (13), pregnant women in LMICs also have access to herbal medicines (14).

Reducing the risk of harm from medication misuse is a global policy priority (15). Global policy documents on antibiotic stewardship, including the WHO’s Antimicrobial Stewardship toolkit (16) focus on prescribing (e.g., monitoring pharmacy dispensing data; hospital drug purchase data; nursing chart administrative data (paper); electronic drug administrative data; e-prescribing records), and do not offer much practical guidance on how to manage self-medication (e.g., use of antibiotics purchased over the counter, or provided by a family relative) (16). Developing and implementing policies and formal procedures that effectively address maternal self-medication in LMICs requires insights and commitment from health professionals (17).

While medical doctors have significant dominance over health care policy in LMICs, including antimicrobial stewardship (18) their views on antibiotics misuse is not well understood. Thus, there has been a growing body of research to better understand and improve how health professionals perceive and manage medication use in patients (19–21). How physicians, a major stakeholder group, view the problem of antibiotic self-medication during pregnancy, especially in LMICs, is an important scientific question to consider.

LMICs often lack clear guidelines for managing antibiotic misuse in pregnant women (22), and existing studies on antimicrobial misuse during pregnancy have primarily assessed *patient* data (23–25), including qualitative research (26). Doctors can offer unique insights on maternal self-medication with antibiotics that go beyond the patient-focused feedback documented in previous studies (10, 23, 24, 26), and help train, and support health professionals to improve antimicrobial stewardship in maternity care globally. Although previous research has examined the views of patients regarding medication use (27–30), including qualitative studies (31–33), doctors’ perspectives have rarely been examined. The few existing studies targeting health professionals were not specific to pregnant women living in a LMIC (19, 20, 30).

The high prevalence of antibiotic self-medication during pregnancy in LMICs (10, 24) is aggravated by inadequate antimicrobial stewardship policies (17). Given the link between antibiotic use and birth complications (34, 35), doctors can offer unique insights on maternal antimicrobial self-medication that help improve patient outcomes (10, 23, 24, 26), and support antibiotic stewardship in maternity care settings. Since medical doctors have considerable influence on health care policy in developing regions, including antimicrobial stewardship (18), we sought to address the following research question; *what are medical doctor’s perspectives on antibiotic self-medication during pregnancy in a LMIC?* To adequately address this question, it is important to capture both views and sentiments. Whereas the former typically depicts opinion and judgement, sentiments convey emotional tone (i.e., ‘positive’, ‘neutral’ or ‘negative’ outlook) (36–38), and can be a significant factor in patient care, clinical decisions, and policy making (39–41). We engaged physicians across three tertiary hospitals in a LMIC, notably staff at the O&G departments. Our goal was to help advance the SMI agenda by providing evidence-based insights that can be used to tailor local policy initiatives on antimicrobial stewardship during pregnancy (10), to ensure they adequately address physician’s views and sentiments, and help identify solutions adaptable to the local health system and context (42).

## Materials and Methods

### Ethics statement

Ethical approval was obtained from the appropriate UK University Research Ethics Committee (LJMU UREC, minimal risk registration number - 22/PSY/066). Ethics approval was also granted by the tertiary hospitals. Written informed consent was obtained from all subjects prior to participation.

### Sample recruitment

Participants comprised 25 medical doctors recruited from three university teaching hospitals in Nigeria, one located in the Southeast, and the other hospitals based in the northern part of the region. Recruitment was implemented by the head of the O&G departments at each hospital, who consulted with colleagues, and then forwarded a list of 147 medical staff available to be interviewed, including 131 medical doctors (see Figure 1). Of this number, 98 doctors were invited for interview, 26 responded, and 25 (17% of the original staff list) were interviewed. A majority of the sample (83.33%) was male. The final sample size of 25 was based on several different justifications: (a) saturation, (b) pragmatic considerations, (c) richness, and volume of data, and (d) threats from sample size insufficiency (e.g., generalisability) (43).

**Figure 1.**
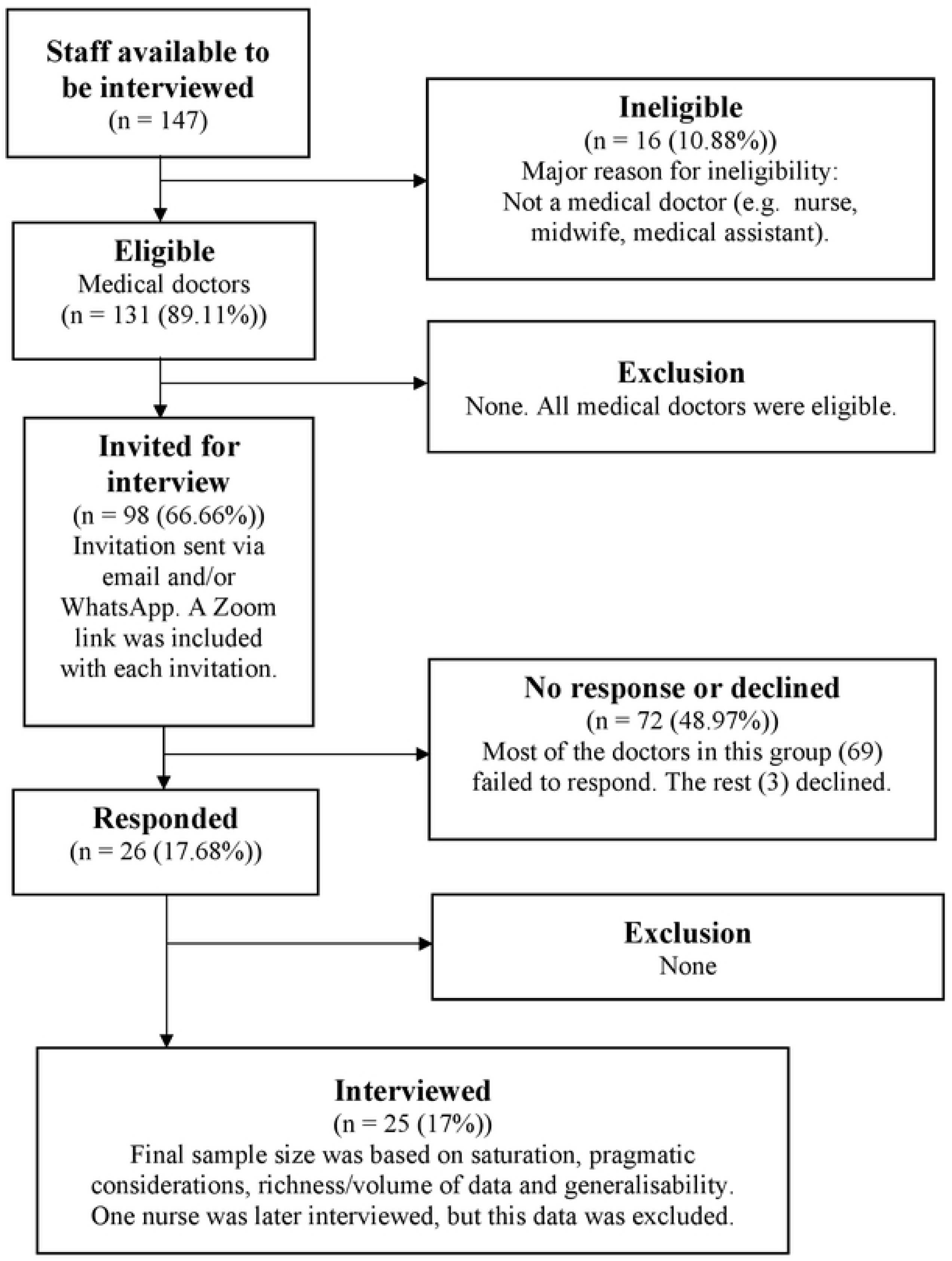
Flow diagram showing recruitment process.

Firstly, the saturation process was based on a subjective criterion: new information threshold (44). By the 25^th^ interview, no (0%) novel information was emerging from the data, beyond what had already been learned from initial earlier interviews. Pragmatic considerations included time constraints imposed by the fix-term contract of the research assistant responsible for transcribing interviews (45). Furthermore, the 25 interviews produced data with sufficient richness and volume for meaningful analyses, with close to (or over) 2000 words generated per interview. Regarding threats from sample size insufficiency, our primary concern was generalisability (43). Although we aimed to recruit from three hospitals located in ethnically diverse regions, allowing some degree of nomothetic generalisability (i.e., potential to draw inferences from the sample to the broader population of doctors), in addition to the more conventional idiographic approach used in qualitative research (capturing the unique individual experiences of the doctors) (46), most interviewees (20 (83.33%)) were ultimately recruited from the south-eastern hospital.

### Data collection

All interviews were conducted remotely, using Zoom Video Communications (Inc) software (47). The interviewer wore headphones (Logitech USB Headset Stereo H570e), with a built-in microphone, for best audio quality. To standardise the interviews each participant was asked fourteen core semi-structured questions, allowing new ideas to be explored further. Some examples of these questions are presented in Table 1. The precise wording for some questions was modified slightly by the interviewer, for more clarity, and/or better pronunciation.

**Table 1.**
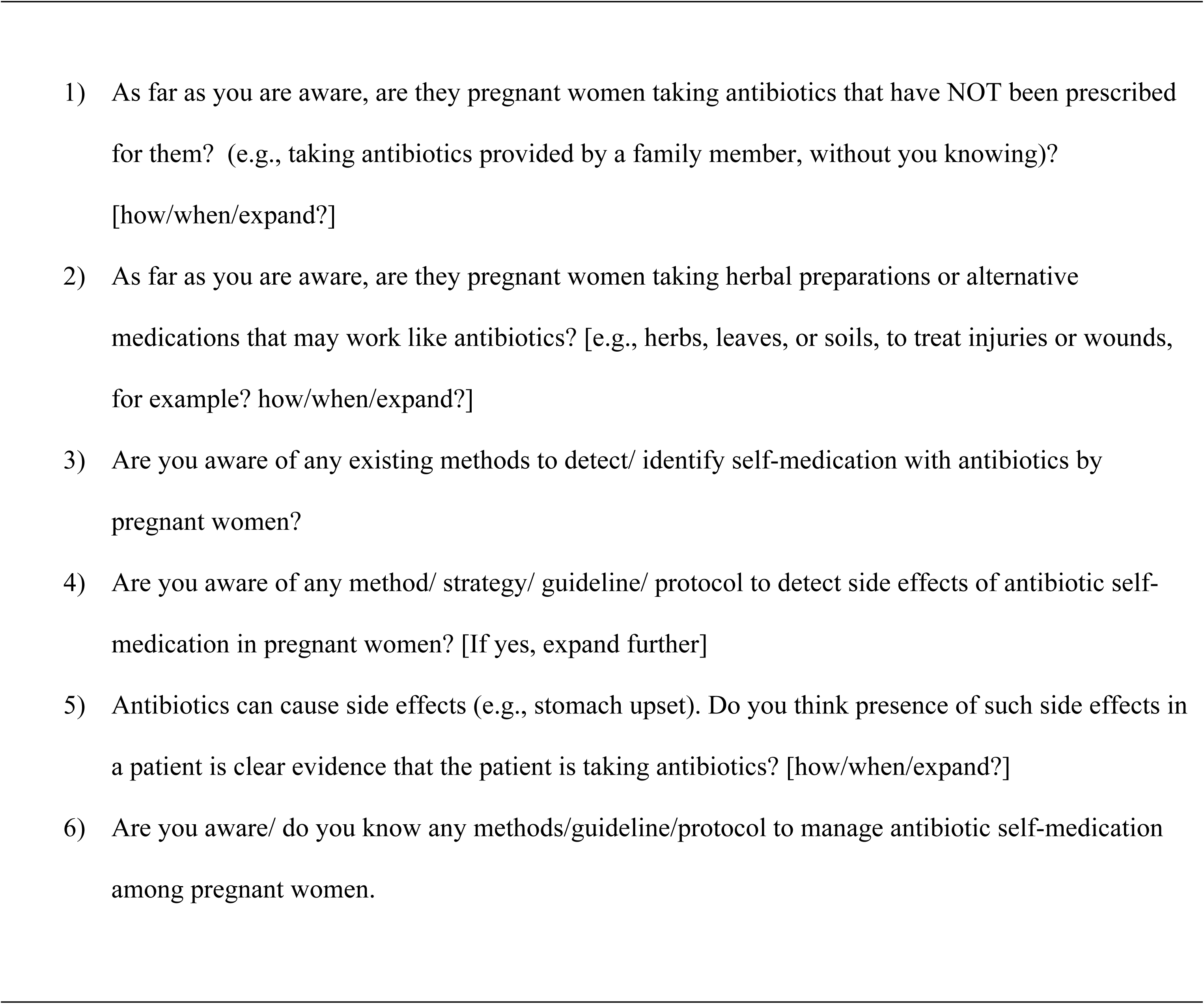
Selection of interview questions.

Several days prior, and again on the day of the interview, the interviewee was sent a Zoom link (via email or WhatsApp), to join the meeting. At the beginning of a session the interviewer clicked the recording button, activating Zoom cloud recording, which comes standard with all Zoom accounts, and allows recordings to be viewed, shared, and downloaded later. The interview was then conducted. Participants had been sent a participant information sheet and consent form prior to the session. Before beginning the interview, the interviewer first obtained oral consent. If the interviewee had not completed and returned their consent form, a new consent form was displayed via screen sharing, and completed by the interviewer, on the doctors’ behalf. Participants had the right to terminate the meeting at any point. At the end of the session, the recording was automatically saved as both an MP4 (video and audio) and MV4 (audio only) file.

On average, each interview lasted about 20 to 40 minutes. Digital file sizes for interview recordings ranged from 12MB to 247.8MB for MP4 recordings, and 3.29MB to 31.2MB for M4A files. The wide disparity in file sizes partly reflects delays due to technical glitches experienced by the doctors, including faulty microphone or speaker setup, slow or unreliable internet connection, lost connectivity, delay in joining the session, and other interruptions.

Interviewees facing repeated or severe technical issues were asked to disable their camera/video function, so the interview was conducted using only audio. Some glitchy sessions ended prematurely, due to lost connectivity, or were terminated by the interviewee. In both cases, the interviewee was invited to reconnect. The interviewer often needed to repeat questions, due to differences in accents and/or poor audio, prolonging the duration of the interview. Overall data collection took approximately 7 months (from February to August 2023). Following data analysis, three participants were contacted via email, to provide feedback on emerging themes.

### Data analysis

Thematic analysis was used to explore themes as it avoided the constraints of more structured analytic methods, such as grounded theory (48). We first adopted a six-phase analytic approach (49). The phases were implemented in a linear fashion, albeit with some overlap, and moving back and forth to verify codes or themes. Using step-by-step guidelines from the literature, this thematic analytic process was further adapted with two additional steps, to create a conceptual model (50).

A research assistant transcribed the audio data to text. The research team shared emails on how best to code the data, including initial coding, to identify distinct concepts, ideas, or topics in each line of text, highlighting lines of text in different colours (each colour capturing a particular concept/code), listing codes in a separate column, and creating categories. Selected interview recordings and their transcripts were later reviewed by a researcher not originally involved in transcribing the data, to ensure transcripts correctly captured the interview recordings. As this study aimed to address a specific research topic – medical doctor’s perspectives on antibiotic misuse during pregnancy – thematic analysis was conducted using a *theoretical* (top-down, or deductive) rather than *inductive* (bottom up) approach. Thus, rather than coding every line of data, we sometimes coded only segments of data that captured something interesting about the research topic (50). Three researchers (HK, KU, RS) independently coded separate batches of interview transcripts. They examined the codes generated from their batch of interview transcripts, to see if they fitted together into themes. Again, this process was completed independently.

The validity of emerging themes was confirmed by additional checks. Two researchers (HK, KU) independently reviewed codes and themes identified, to determine if they made sense, and resolve any queries. They read interviewee statements associated with each code and/or theme and considered whether the data really did support it. Six specific issues were considered (see Table 2). Final definition of themes was based on their content, with our focus on the ‘essence’ of each theme, and potential for overlap and/or presence of subthemes (53). The primary determination was that potential subthemes (e.g., ‘side effects’, ‘herbal self-medication’) had sufficient data and unique content, to be classed as major themes. Also, there was no definitive overarching theme. We used a psychological framework applied to medication use as the basis for defining and interpreting emerging themes and any underlying concepts (50). We reviewed the work of others (51) to help conceptualise themes, with the goal of addressing the research question, and underscoring the study’s contribution to knowledge (50). There was ongoing reflection and engagement with the data, including revisiting concepts.

**Table 2.** Selected issues highlighted during independent review of codes and themes.

| Issues considered | Reviewer feedback |
| --- | --- |
| 1. Do the themes make sense? | Feedback from both reviewers suggested the themes made sense. |
| 2. Does the data support the themes? | Reviewer feedback indicated agreement that the data supported emerging themes. |
| 3. Am I trying to fit too much into a theme? | 'Detection' as a concept incorporated the largest number of codes, per interview, but the underlying text was relevant. |
| 4. If themes overlap, are they really separate themes? | The concepts 'use of herbal medicines' and 'self-medication' partly overlapped, but distinct content and volume of data justified separate themes. |
| 5. Are there themes within themes (subthemes)? | Reviewers agreed no subthemes materialised. 'Guidelines' were occasionally referenced within other themes but emerged a dominant facet across the dataset. |
| 6. Are there other themes within the data? | Two additional concepts emerged (e.g., 'cultural mindsets'), but each was attributable to just one interviewee. |

The researchers performed several robustness checks: reviewing records of over 800 email conversations (spanning approximately 10 months (from February to November, 2023), between members of the research team, on the data collection and analysis process, to help track our decision making, and ensure consistency (e.g., ensuring codes/themes generated by one researcher are reviewed by a different colleague) (52); considering negative or conflicting perspectives by examining quotations that seemed to contradict emerging themes (53); conducting triangulation, by comparing the data with results from previous studies using a quantitative approach (54); checking for clarity of thought, by examining initial suppositions documented in our initial research proposal (application for research funding), to reflect on how early presumptions converged with or diverged from emerging themes (55); conducting respondent validation, whereby the head of one O&G department checked the accuracy of themes, to identify any issues that need correction and/or elaboration and determine whether our conclusions reflect their experiences (52).

We used Boardflare’s analytic software for text in Excel spreadsheets to analyse participants ‘sentiments (56). The analysis is performed locally using a pretrained open-source transformer machine learning (ML) model for natural language processing (NLP): we selected the Twitter-roBERTa-base for Sentiment Analysis model, which generates ‘Positive’, ‘Negative’, and ‘Neutral’ output labels for each word or sentence, together with a confidence score (57). This model has been pretrained on data containing 124 million twitter messages. This content generally encapsulates unformatted, informal and colloquial language, similar to everyday conversation, and hence is especially suited to text that includes unrehearsed, casual, and grammatically flawed speech, slang and other forms of spontaneous ‘everyday’ language (58). We opted for ‘document-level’ rather than ‘sentence-level’ sentiment analysis: the former assesses any sentiment-bearing text, whereas the latter evaluates sentences (59). Although sentence-level can provide useful insights, it may introduce additional errors and complexities, notably ‘overfitting’ (the model detects artificial patterns in the sentence that don’t reflect the prevailing sentiment) and risk of misinterpretation (the model misunderstands sentences, resulting in sentiment labels) (60), especially given the easily misunderstood linguistic ambiguities present the current interview transcripts (61).

ChatGPT assisted with data cleaning: a bespoke set of AI instructions (inputs) were used to remove unsuitable text, such as formatting inconsistencies, and irrelevant information, thereby improving the performance of the text analytic model (62). For example, ChatGPT was instructed to remove unwanted abbreviations, sentences starting with the word ‘interviewer’, and interviewer phrases describing undecipherable interviewee statements (e.g., ‘unclear speech’, ‘mumbled speech’ or ‘overlapping speech’), or technological irregularities (e.g., ‘signal distortion’). Where appropriate, these AI instructions were customised and/or tailored to individual transcripts, to address any unique or unusual transcript-specific phrases, or grammatical distortions. For example, the identity of each interviewee (represented by a code on their transcript) was unique, and ChatGPT had to be specifically instructed to remove each distinct identifier. We did not extract nouns and adjectival phrases because these were deemed to relate directly to the discussion, and sentiments expressed (e.g., the noun ‘patient’ refers to a person who self-medicates during pregnancy, which in turn may convey a certain sentiment) (58). In addition to using ChatGPT for data cleaning, almost all transcripts had to be further edited manually, to remove unrelated or irrelevant text (e.g., discussing consent or reimbursement procedures). A One-Way Chi-square test and One-Sample Wald-Wolfowitz Runs test for randomness were used to analyse the sentiment categories generated from Boardflare’s ML/NLP software. The analysis was performed separately on data from each interview transcript. We applied a Bonferroni correction (*p* < 0.002), to help control for increased type 1 error rates.

## Results

### Perspectives on maternal antimicrobial self-medication

Seven major themes emerged depicting three broad opinions relating to antibiotic treatment, patient behaviour, and clinical practice guidelines. These domains are summarised in Table 3.

**Table 3.**
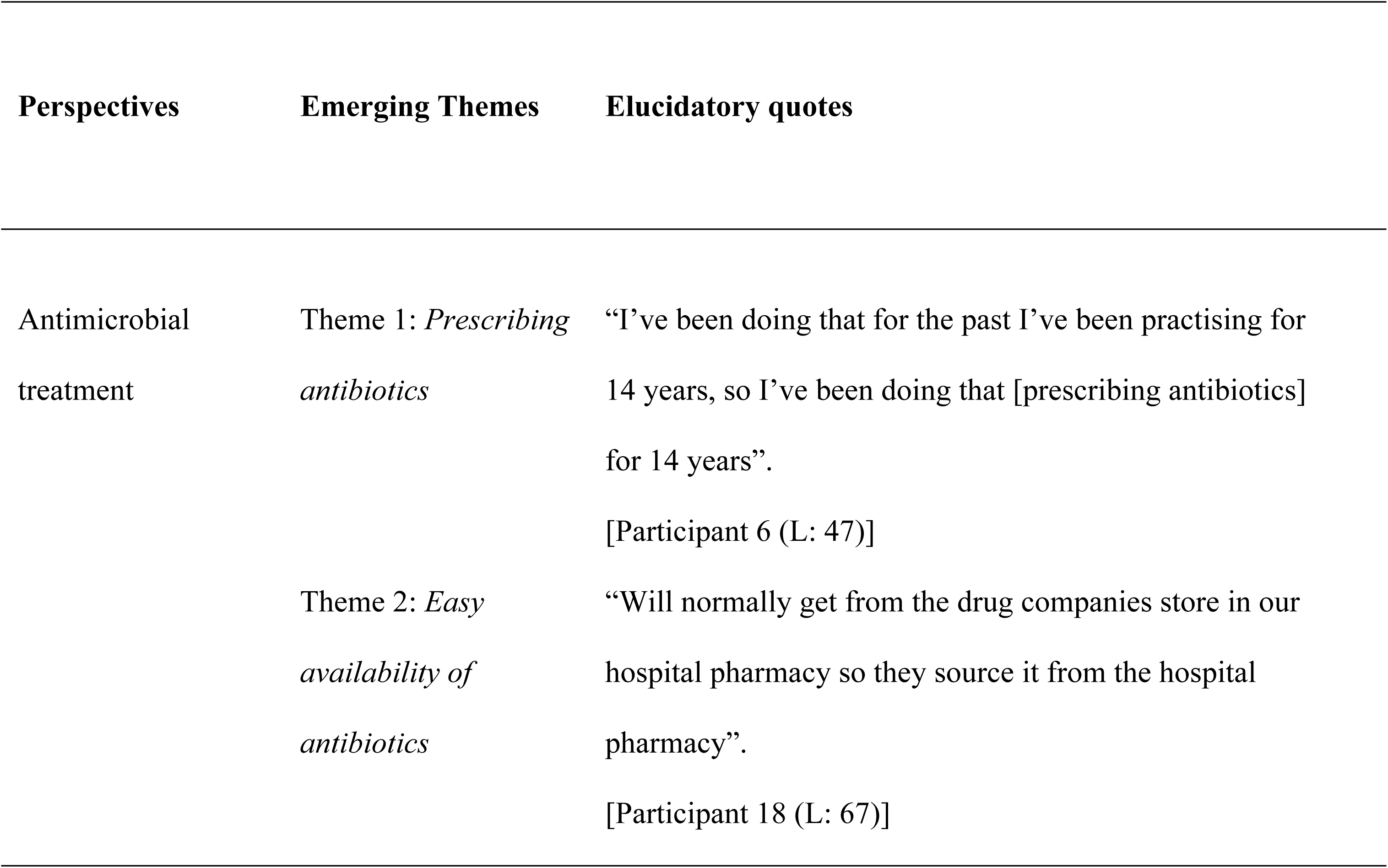
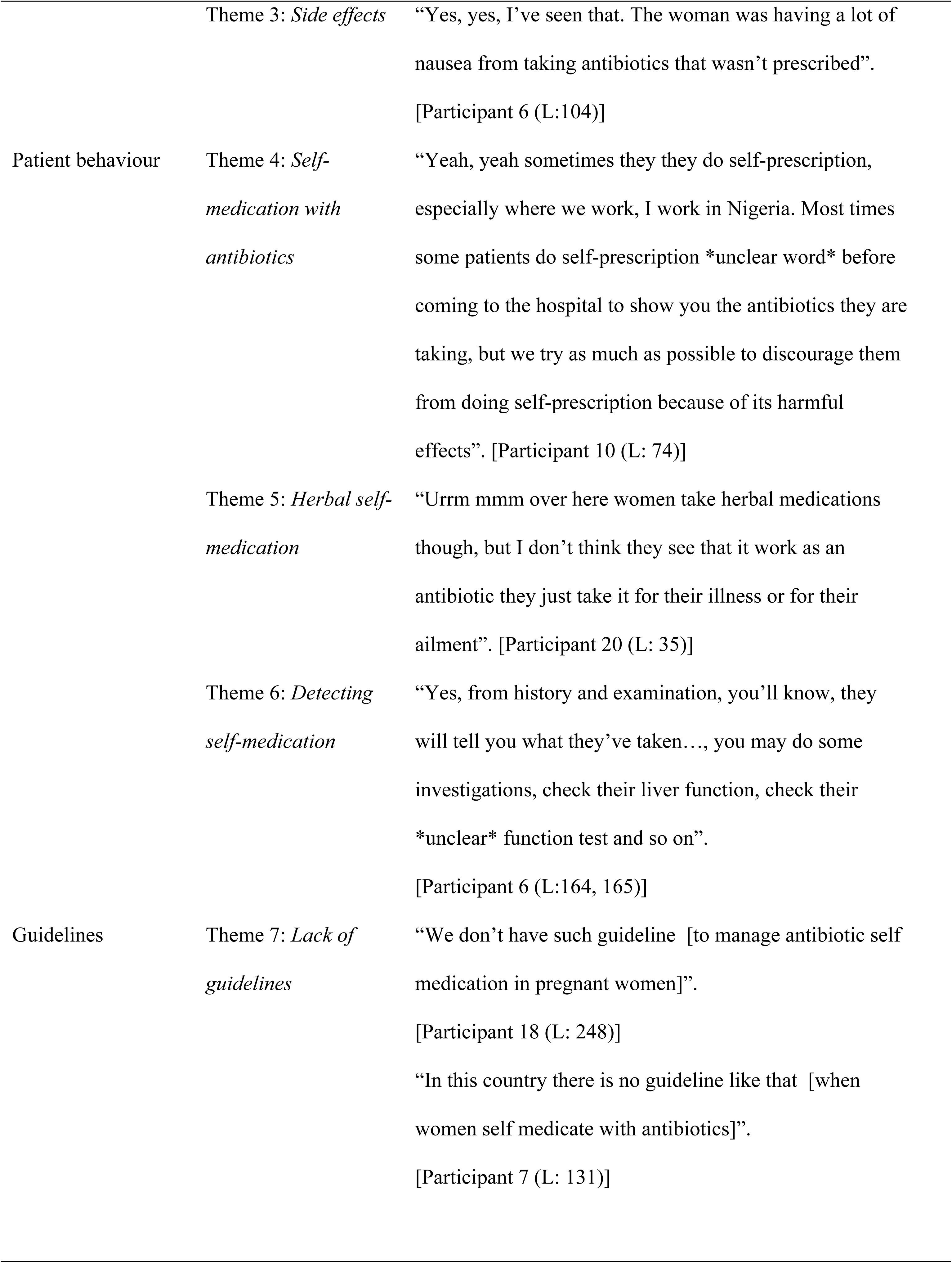
Emerging themes and descriptions.

| Perspectives | Emerging Themes | Elucidatory quotes |
| --- | --- | --- |
| Antimicrobial treatment | Theme 1: *Prescribing antibiotics* | “I’ve been doing that for the past I’ve been practising for 14 years, so I’ve been doing that [prescribing antibiotics] for 14 years”.<br>[Participant 6 (L: 47)] |
|  | Theme 2: *Easy availability of antibiotics* | “Will normally get from the drug companies store in our hospital pharmacy so they source it from the hospital pharmacy”.<br>[Participant 18 (L: 67)] |
|  | Theme 3: *Side effects* | “Yes, yes, I’ve seen that. The woman was having a lot of nausea from taking antibiotics that wasn’t prescribed”.<br>[Participant 6 (L:104)] |
| Patient behaviour | Theme 4: *Self-medication with antibiotics* | “Yeah, yeah sometimes they they do self-prescription, especially where we work, I work in Nigeria. Most times some patients do self-prescription *unclear word* before coming to the hospital to show you the antibiotics they are taking, but we try as much as possible to discourage them from doing self-prescription because of its harmful effects”. [Participant 10 (L: 74)] |
|  | Theme 5: *Herbal self-medication* | “Urrm mmm over here women take herbal medications though, but I don’t think they see that it work as an antibiotic they just take it for their illness or for their ailment”. [Participant 20 (L: 35)] |
|  | Theme 6: *Detecting self-medication* | “Yes, from history and examination, you’ll know, they will tell you what they’ve taken..., you may do some investigations, check their liver function, check their *unclear* function test and so on”.<br>[Participant 6 (L:164, 165)] |
| Guidelines | Theme 7: *Lack of guidelines* | “We don’t have such guideline [to manage antibiotic self medication in pregnant women]”.<br>[Participant 18 (L: 248)]<br>“In this country there is no guideline like that [when women self medicate with antibiotics]”.<br>[Participant 7 (L: 131)] |

#### Views on antimicrobial treatment

Antibiotic prescribing was a key aspect of antenatal care, with most doctors reporting many years of clinical experience. There was some variability regarding the frequency of prescribing, years of experience prescribing, distribution of antibiotics, setting in which antibiotics are prescribed, and the medical conditions antibiotics are prescribed for, including vaginal discharge, urinary tract infections, and upper respiratory tract infection.

> ***“Yes, occasionally when they have urm *unclear word* vaginal discharge *unclear speech* vaginal discharge and so prescribe antibiotics for *unclear word*”.***
>
> [Participant 12 (L: 38)]

Participants noted the easy availability of antibiotics, remarking that pregnant women can obtain antimicrobial drugs from multiple sources, including the hospital pharmacy and over-the-counter drug vendors in the community.

> ***“Mostly over the counter drug erm pharmacies um small business um drug vendors *unclear word*, I would say um it’s not as restricted, so they tend to get It a bit more freely than you think”*.**
>
> [Participant 11 (L: 73)]

Participants highlighted the possible side effects of taking antibiotics, including non-prescribed medication. These adverse reactions included congenital abnormalities and other pregnancy complications. However, most doctors noted that presence of side effects alone was not sufficient proof that a patient was self-medicating with antibiotics. Further clinical assessment was necessary, including history taking, to detect the cause(s) of side effects.

> ***“Erm there’s some women actually that we know that use drugs in early pregnancy and they have malformed foetus’s babies delivered preterm or sometimes they have miscarriage and then some obvious abnormality…, But then it’s difficult to pin down on the real cause of this is result of antibiotics”.***
>
> [Participant 15a (L:208, 210)]

#### Views on patient behaviour

Self-medication with antibiotics during pregnancy was seen as a common practice, with several doctors describing it as an accepted practice they try to discourage. Various motivating factors were identified including pressure or encouragement from friends or family to take antibiotics and needing to alleviate illness symptoms.

> ***“Urm it happens maybe urm with friends might have taken some antibiotics and urm will tell the other ones that okay I had similar problem this what I took”***.
>
> [Participant 14 (L: 57)]

Participants believed pregnant women often used herbal medicines, with some doctors discouraging the practice. Although one interviewee felt herbal usage was rare, most participants suggested it was a common practice amongst pregnant women. Different types of locally prepared herbs, often unnamed, were used. These products were not necessarily treated as an alternative to antibiotics, but rather simply employed to treat illness symptoms. The high cost of medicines was considered a motivating factor.

> ***“Yes, we have a very large urrr local traditional population that actually prefer herbal medications some of them don’t prefer herbal medication but because of costs of *unclear word* medicines they *unclear word* go with urr they have urr some of those traditional preparations”.***
>
> [Participant 25 (L: 67)]

Many doctors relied on their clinical experience, direct questioning, and/or history taking, to determine if a pregnant woman was taking unprescribed antibiotics. Nevertheless, most were receptive to the idea of a practical diagnostic point-of-care test for detecting self-medication, for example a questionnaire and/or technological device. It was critical that such a test was portable, affordable, and did not require electricity.

> ***“Yes, the dangers are many so it would be good to have such a test. So that such women were identified they would be counselled and not have bad side effects or adverse effects and reduce morbidity and mortality rates. So, it would be good to have such tests…., Yes, something that is portable would be good, a point of care test…., it would not require electricity, that would be good”*.**
>
> [Participant 1 (L:122, 126, 128)]

#### Views on guidelines

Some participants expressed a lack of clear policy guidance on how to manage self-medication during pregnancy. Although interviewees did mention various guidelines used in antenatal care, including labour ward and drug prescribing protocols, they were not aware of any regulations specific to antimicrobial self-medication. Some doctors referenced foreign standards, for example RCOG (Royal College of Obstetrics and Gynaecology) guidelines, but not in relation to self-medication. This recurring theme did not appear to overlap convincingly with any illness representation from the CSM.

> ***“No, I cannot say for now. There are no guideline, I am aware there is a guideline on safety of drugs, categories of drugs used in pregnancy by the *unclear word* CDC, categories, category 1, 2, 3. That one I am aware, but guidelines no, I am not aware of any guideline”.***
>
> [Participant 1 (L:131)]
>
> ***“mmmm ummm no I’m not aware of any guideline…, for managing antibiotic um side effects of um self-medication…, okay I’ve not actually come across that”.***
>
> [Participant 20 (L:107, 111, 113)]

### Sentiments towards maternal antimicrobial self-medication

In total we analysed 1484 lines of text, ranging from single word/responses to full phrases and sentences, using Boardflare’s analytic software in Excel (56). This analysis generated a column of sentiment labels (‘negative’, ‘neutral’, ‘positive’) corresponding to lines of text from each interview transcript. We then used a One-Sample Chi-square test to compare the combined total of all observed sentiment categories to the null hypothesis (i.e., labels occur with equal probabilities). This analysis revealed a distinct pattern, whereby the vast majority (77.7%) of sentiment categories were ‘neutral’, compared with ‘negative’ (12%) and ‘positive’ (10.3%) labels, (χ2 (1, N = 1484) = 1314.858, *p* < 0.001). Figure 2 illustrates the distribution of sentiments for all lines of text combined. To check if the sequence of sentiment categories is random or denotes a pattern, we performed a One-Sample Wald-Wolfowitz Runs test for randomness. To conduct this test, we organised the sentiment labels into binary data (neutral/positive; neutral/negative; positive/negative).

**Figure 2.**
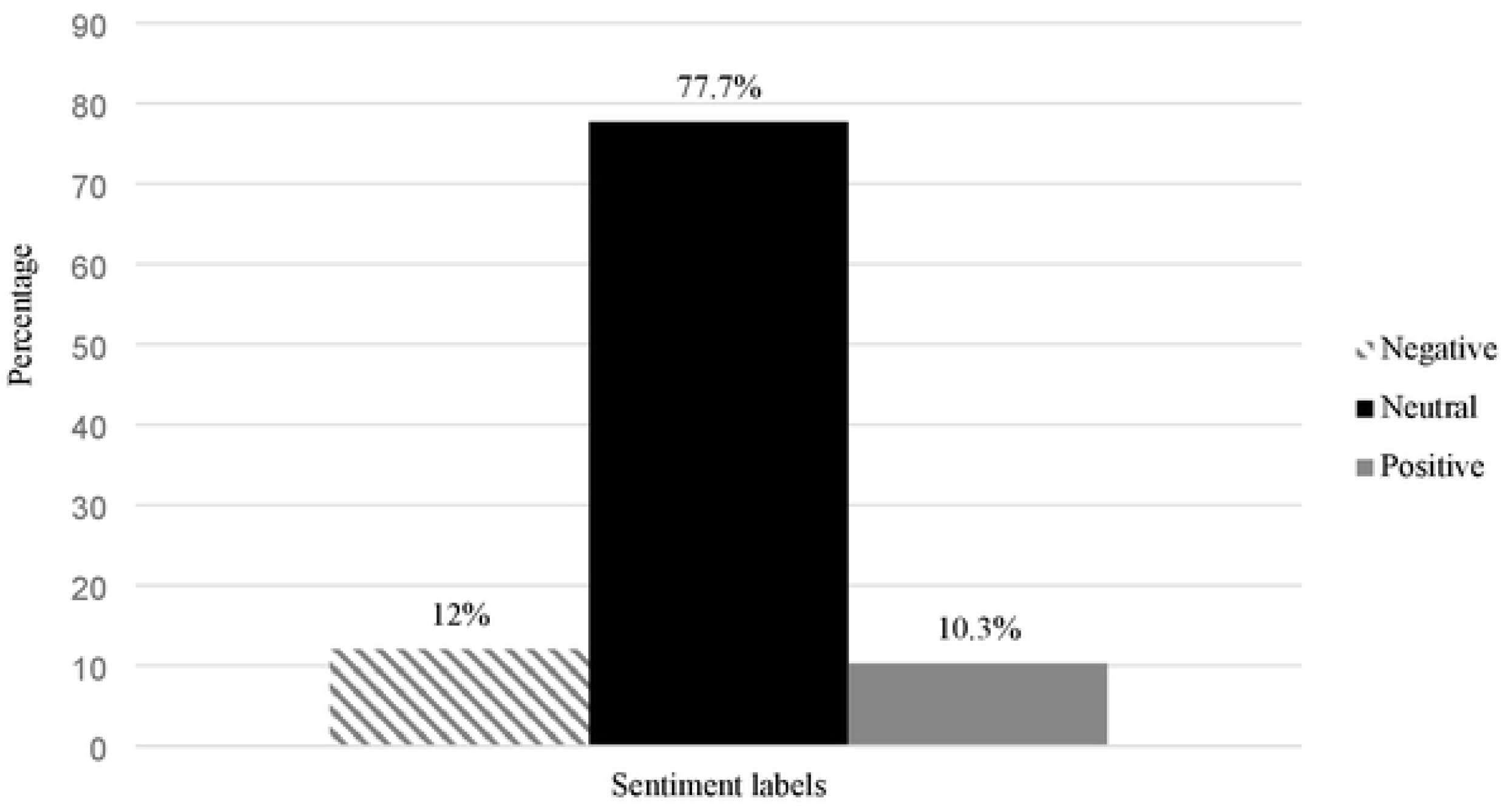
Bar chart showing total distribution of sentiment labels across all transcripts.

The observed patterns were significantly different from what would be predicted under the null hypothesis of randomness (see Table 4), depicting an abnormally high rate of ‘neutral’ labels, compared with ‘positive’ (*z* = −36.058, *p* < 0.001) and ‘negative’ (*z* = - 36.410, *p* < 0.001) categories. There were also significantly more ‘negative’ compared with ‘positive’ labels (*z* = −18.111, *p* < 0.001), from what can be expected randomly.

**Table 4.**
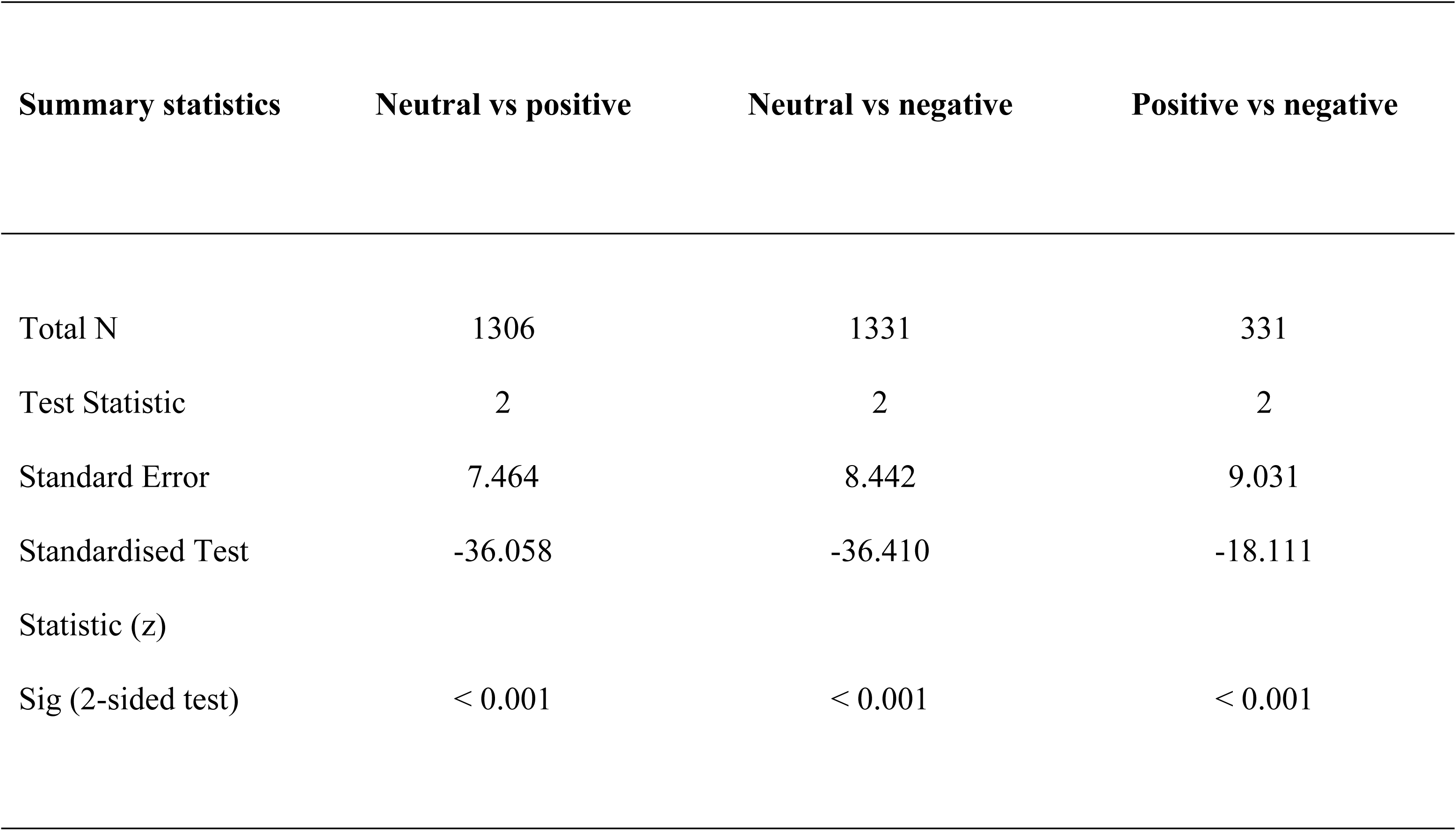
Summary of Wald-Wolfowitz Runs test for randomness of sentiment labels.

## Discussion

This is the first study to explore the opinions and sentiments of medical doctors from a LMIC regarding maternal self-medication with antibiotics, using both qualitative and sentiment analysis. Although past studies have examined health professionals’ views on patient’s medication use (19, 20, 30), there has been limited research relating specifically to pregnant women. We found that the doctors expressed views regarding patient behaviour, antimicrobial treatment, and clinical practice guidelines, but also displayed a predominantly neutral sentiment towards the topic of discussion. Overall, these findings extend previous research on medication misuse in pregnant women from LMICs, particularly Sub-Saharan Africa (10, 14, 23–26).

Self-medication, especially with medicinal herbs, was a significant concern, supporting previous research showing the practice is widespread in LMICs (63–65), often to relieve illness symptoms (66). The doctors believed use of herbal medicines during pregnancy is endemic amongst pregnant women (23, 24, 26). Although there was no consensus regarding the type and/or biochemical content of herbs used, limited access to antibiotics (e.g., due to high financial cost) was believed to be a factor encouraging reliance on traditional medicines. While certain herbs do have antimicrobial properties, a typical medicinal herb (e.g., plant extract) may contain over 150 chemical ingredients, making identification of adverse effects and drug interactions difficult (67). Thus, the doctors did not recommend herbal medicinal use during pregnancy (68).

Detecting and monitoring medication misuse was a challenge. While many doctors relied on history taking, and/or recording side effects that might indicate antibiotic use, most accepted the need for a cost-effective and practical point-of-care diagnostic tool for detecting self-medication in antenatal care settings. Although doctors in Western countries can measure the level of antibiotics in the blood, using a rapid blood test (69–72), this procedure is invasive and requires expensive biomedical equipment and materials (e.g., biosensors and clinical microbiology laboratories) which may be unaffordable in poor countries (73–75). Thus, what is needed is an affordable test that can be easily administered, with minimal technological or resource requirements. Pregnant women living in LMICs often rely on resource-deprived primary healthcare centres lacking the staff, equipment, and tools to deploy expensive resource-intensive diagnostic tools (76).

Antibiotics as a form of antimicrobial treatment was a distinct topic. The ease with which pregnant women obtain antibiotics without a prescription (i.e., the availability of antibiotics) was particular concern. Previous research has shown that medicinal drugs are readily available in LMICs, (e.g., from local traditional healers, family members, or over-the-counter sales) (6, 23, 77). Naturally occurring herbal products are also easily accessible (14): they can be obtained from multiple sources, including friends and family, or local traditional healers (native doctors) (64, 78, 79). Finally, the doctors discussed the potential side effects of antibiotic misuse during pregnancy, which also reflects treatment beliefs (80). Their view that the presence of side effects alone did not demonstrate self-medication seemed to depict an interesting conceptual domain, perhaps unique to health professionals, given that the average patient is unlikely to be familiar with the pharmacology of antimicrobial side effects (81).

Lack of clinical practice guidelines specific to self-medication during pregnancy was underscored. This view seemed to encapsulate organisational rather than individual-level themes (e.g., workplace rules, procedures, standards). Current antibiotic stewardship policy focuses on prescribing (16), and there is limited policy guidance on how to manage self-medication (22), including methods for detecting antimicrobial misuse. These policy gaps are evident in WHO policy documents on antibiotic use in LMICs, including the WHO’s Antimicrobial Stewardship toolkit (16). Although some doctors relied on foreign guidelines on antimicrobial stewardship (e.g., RCOG), procedures specific to a particular geographical context may be necessary, given resource challenges, cultural considerations, and other societal constraints unique to a particular region (17).

The importance of sentiment in clinical settings is well documented (38–41). The dominance of apparently neutral sentiment observed here may have both unfavourable and beneficial interpretations. On the one hand, it may signal that self-medication during pregnancy isn’t viewed by local doctors as an especially important or noteworthy concern requiring urgent action (36, 37). This presumed apathy seems to echo published literature highlighting a lack of serious initiatives to address maternal self-medication in many LMICs (10). A lack of urgency may have both clinical and policy implications, for example affecting the exigency with which medical doctors discuss the issue with patients, and/or lobby for clearer guidelines on antimicrobial stewardship (40). On the other hand, neutral sentiment may suggest doctors understand and are familiar with the problem of maternal self-medication, for example use of herbal medicines (10, 82), and hence do feel the need to give undue attention to it (37). Furthermore, a neutral demeanour may also imply a rational (non-emotional) approach to the problem, for example in how they view their interactions with patients or manage side effects (36).

Negative sentiments were sparse but noteworthy, often alluded to themes already documented in the literature, such as common illnesses for which antibiotics need to be prescribed (e.g., urinary or respiratory tract infections) (83), the seriousness of self-medication (10), side effects of antibiotic misuse (e.g., rashes) (84), uncertainty about ingredients contained in medicinal herbs (82), lack of effective methods for detecting self-medication (85), and/or the need for specific guidelines on managing antimicrobial self-medication (86). Positive demeanour, albeit more sporadic compared with negative sentiment, often signalled endorsement of the idea of a non-invasive tool or test to detect maternal self-medication, despite the challenges (85). The preponderance of negative labels, relative to positive sentiment, is a pattern that may have wider implications for patient engagement and necessitates further inquiry (87).

### Strengths and limitations

This study has several limitations. The sample size and composition are problematic as we only interviewed a limited number of medical doctors, mostly from one tertiary hospital. Furthermore, while doctors are a key stakeholder group, exerting considerable influence on health care delivery (18), research suggests most pregnant women in LMICs, particularly those living in rural areas, have limited access to a doctor (88). Over two-thirds (72%) of pregnant women are cared for by a worker without formal medical training, such as a traditional birth assistant (89). Many pregnant women only visit a health facility (e.g., a rural clinic) in an emergency (88), suggesting medical doctors may have limited opportunity to thoroughly investigate medication misuse. Women who do consult with a doctor are unlikely to share their self-medication habits, due to concerns about disapproval, and other cultural factors (64, 77, 90). Thus, future research needs to interview other stakeholder groups with unique insights on self-medication in pregnancy.

Another possible limitation is the use of a deductive (top-down) approach to thematic analysis, driven by the research *topic*. It is possible that an inductive (bottom-up) approach based primarily on the data, rather than a preconceived research question, would have generated novel constructs not captured by the (rather more conceptually restricted) deductive approach. For example, two minor concepts emerged that didn’t evolve into major themes – one referencing established cultural mindsets on medication use, while the other highlighted the role of independent organisations who supply antibiotics without the need for a doctor’s prescription. It is possible these ideas might have evolved into major themes had we used an inductive approach, unconstrained by an a priori focus on specific narratives.

The limitations of sentiment analysis using ML/NLP models has been well documented, including an inability to understand context, or detect irony or sarcasm (58, 60, 62). Interviews often used ambiguous context-dependent language containing slang, linguistic quirks, and cultural nuances that contribute to inaccurate interpretations. For example, the meaning of (and sentiments associated with) the term ‘self-medication’ in relation to medicinal herbs may be strongly dependent on context, since some African doctors view patient use of herbal medicines as acceptable (91). Such ambiguity can confuse sentiment analysis models and leads to errors in interpretation. Another problem is that most ML/NLP models are trained on conventional English text (57), meaning the accuracy of sentiment labels will be lower for words, phrases or expressions that are not standard or widely recognised (92).

Despite these constraints, the robustness of our findings was confirmed through a series of checks for methodological rigour (Noble & Smith, 2015). First, although the limited sample size has implications for data interpretation, we have been transparent on sample selection, and the methodological justifications used, including saturation, pragmatic considerations, richness, and volume of data, and sample size insufficiency (e.g., generalisability) (43). While recruitment was mostly from one hospital, negating nomothetic generalisability, the data still offers idiographic insights (i.e., captures the unique individual experiences of the doctors), as expected in qualitative research (46).

Second, we conducted meticulous record keeping, in the form of hundreds of email conversations between members of the research team, which documented data collection and analytic procedures. This provided a clear decision trail, ensuring our methods were consistent, and transparent (52). All interviews were recorded using the same software (e.g., Zoom), and stored in the same location (cloud recording), and in similar formats (MP4 and MV4). To enhance accuracy in data interpretation, selected interview recordings, and their transcripts, were reviewed by a researcher not originally involved in transcribing the information, to ensure the recorded dialogue was accurately captured in the transcripts. Third, we examined differences across interviewee experiences, to ensure conflicting perspectives are considered (53), including concepts mentioned by just one interviewee (e.g., cultural mindsets), and conflicting accounts within themes (e.g., ambiguity regarding the prevalence of herbal self-medication, and at least one doctor had not encountered patients who self-medicate). Fourth, we performed data triangulation (52), by comparing our findings with outcomes from previous quantitative research (54).This process demonstrated some correspondence, for example the widespread use of medicinal herbs (13).

Fifth, we demonstrated clarity in thought from initial preconceptions to subsequent inferences (55), by assessing how emerging themes and sentiments compared with our initial suppositions documented in the application for research funding. For example, the lack of policy guidance on antibiotic stewardship in pregnancy, a factor contributing to negative sentiment, reflected our initial presuppositions. Sixth, data collection and thematic analysis were conducted by researchers from four different ethnic backgrounds (Nepalese, Jewish, Nigerian, Indian,), helping reduce cultural bias during the process (55). Two researchers conducted independent checks, to verify initial coding, and emerging themes. Finally, we conducted respondent validation, to verify the accuracy of the findings and identify any points that need correcting or clarifying (52).

### Implications

Although the SMI is currently a global priority, with various regional, national, and international strategies implemented to address maternal and child health (1, 2), the implications of maternal self-medication for meeting Safe Motherhood and SDG-3 objectives in LMICs has yet to be addressed (93). Since medical doctors are an influential stakeholder group (18), with influence on health policy (17), their views and sentiments on maternal self-medication are relevant to meeting SDG-3 targets. The current findings encapsulate the scale of the challenge. Firstly, doctors are keenly aware pregnant women self-medicate with antibiotics, suggesting a readiness to engage with any regional or global SMI campaigns to improve medication safety during pregnancy. Secondly, a focus on antimicrobial *prescribing* in tertiary hospitals, combined with a lack of clear policy guidelines for managing *self-medication*, may hamper coordinated campaigns to improve medication safety during pregnancy. Third, lack of practical point-of-care diagnostic tools to detect and monitor maternal self-medication, in a challenging environment, means LMIC doctors often have no way of collecting baseline data, and tracking changes in antibiotic misuse, over time, to gauge the impact of any SMI initiatives promoting safe motherhood (94–97), and the longer-term impact on rates of birth defects, and maternal, neonatal and child mortality (2). LMICs often lack a coordinated and comprehensive surveillance system on medication safety during pregnancy (94–97). Fourth, the dominance of a neutral sentiment may have clinical implications for doctor-patient interactions regarding antimicrobial self-medication, detecting antibiotic misuse in patients, and physicians’ motivation to improve antimicrobial stewardship and/or develop clear protocols (40). As a major stakeholder group, medical doctors are essential for generating regional or international debate on antimicrobial stewardship in pregnancy (17), with particular focus on self-medication (23, 24, 26), so a neutral demeanour (e.g., lack of urgency) may be both problematic and beneficial (36, 37).

## Conclusions

Medication safety in pregnancy is integral to the SMI global agenda and SDG-3 initiatives in LMICs. Although medical doctors living in LMICs are an influential stakeholder group, how they view and feel about the challenge of antimicrobial self-medication during pregnancy has not been well understood. This is the first study to explore this topic in a West African context, using both qualitative and sentiment analysis. We found that views on patient behaviour (e.g., use of herbal medicines), antimicrobial treatment (e.g., antibiotic availability, side effects), and clinical practice guidelines, and prevailing neutral sentiment (suggesting apathy), captured how local doctors regard pregnant women’s use of non-prescribed antibiotics. These findings can be used to tailor local SMI initiatives on antimicrobial stewardship during pregnancy so that they address physician’s concerns and emotional disposition, including the lack of clear guidelines and an apparent lack of urgency.

## Acknowledgements

We would like to thank the Impact Officers at Liverpool John Moores University (Research and Innovation Services) for their support.

## Data availability statement

The interview recordings generated and/or analysed during the current study are not publicly available because individual privacy could be compromised. The data consists of MP4 (video and audio) and MV4 (audio only) files containing personal information, which is related to an identified or identifiable natural person, and hence subject to the protection requirements set out in the Data Protection Act 2018 (the UK’s implementation of the EU’s General Data Protection Regulation (GDPR)). The recordings contain each interviewees full name, phone number, email, signature, race, ethnic origin, and other personal details. Fully anonymised interview transcripts are available from the corresponding author on reasonable request, as personal data that has been anonymised is not subject to the UK GDPR.

## Author contributions

**K Umeh:** Conceptualisation (lead); data curation (lead); formal analysis (lead); funding acquisition (lead); investigation (lead); methodology (lead); project administration (lead); supervision (lead); validation (lead); writing – original draft (lead); writing – review and editing (lead). **S Adaji:** Conceptualisation (lead); investigation (supporting); methodology (supporting); project administration (supporting); supervision (supporting); writing – review and editing (equal). **G.U. Eleje**; Conceptualisation (supporting); methodology (supporting); project administration (supporting). **E.O. Umeh**; methodology (supporting). **S Ushie**; methodology (supporting). **C.G. Okafor**; methodology (supporting). **C.B. Oguejiofor**; methodology (supporting). **S. Bature**; project administration (supporting). **N H Madugu**; project administration (supporting). **U Bawa**; project administration (supporting). **M Sacks**; data curation (supporting); formal analysis (supporting); investigation (supporting); project administration (supporting); supervision (supporting); validation (supporting). **R. Singh**; formal analysis (supporting); investigation (supporting). **H. Karuppuchamy**; formal analysis (supporting); investigation (supporting).

